# Undergraduate medical students in India are underprepared to be the young-taskforce against Covid-19 amid prevalent fears

**DOI:** 10.1101/2020.04.11.20061333

**Authors:** Vishwesh Agarwal, Latika Gupta, Samira Davalbhakta, Durga Misra, Vikas Agarwal, Ashish Goel

**Affiliations:** Mahatma Gandhi Missions Medical College, Navi Mumbai; Department of Clinical Immunology Sanjay Gandhi Postgraduate Institute of Medical Sciences Lucknow, India; Byramjee Jeejeebhoy Medical College, Pune; Department of Medicine, University College of Medical Sciences, New Delhi

**Keywords:** Coronavirus, Undergraduate medical students, India, COVID

## Abstract

**Background:** The healthcare system in India faces an acute shortage of front-line doctors to fight the Covid-19. Thus, the recruitment of undergraduate medical students into the health care force is being considered by many state governments. A survey was conducted amongst undergraduate medical students to understand their knowledge, attitude, and preparedness towards the ongoing pandemic.

**Methods:** An anonymized survey on a cloud-based website (Survey Monkey®) comprising 33 questions was served to medical students.

**Results:** Of 616 (24.6%) respondents among 2500 invitees across six medical schools in India, most (54.1%) were in the final year of their undergraduate training program. Knowledge regarding viral transmission, clinical features, laboratory diagnosis, and drugs being tried out in Covid-19 was adequate among most students. However, understanding of the incubation period 123 (20%) and time to symptoms 30 (4.8%) was less than satisfactory. Three-fourths (75%) were unaware of the treatment guidelines for Covid-19, and one-quarter (155, 25.1%) were unaware of the required precautions during management. Moreover, 179 (29.1%) were unaware that Covid-19 causes an asymptomatic or minor illness in young individuals. Nearly 70% were reluctant to attend clinics from fear of getting infected or passing on to others. Besides, 250 (40.6%) were not updated on Covid-19, and most (486, 78.9%) resorted to social media for information on Covid-19.

**Conclusion:** Prevalent fears and inadequate understanding of Covid-19 suggest that undergraduate medical students are not prepared to be the front-line taskforce in the current pandemic.

## Introduction

The recent COVID19 pandemic has led to near-complete saturation and exhaustion of healthcare resources worldwide, including India. Tapping into the massive human resource from undergraduate medical schools could be a potential solution,^1^ and many state governments in India are considering it. Despite the fact that most graduation school students are still in the learning phase of their career and are neither experienced nor licensed to practice, emergent circumstances may force state governments to engage them in the arduous task of volunteer-force against Covid-19. However, it raises concerns regarding the safety of both patients and students. Therefore, we undertook an online survey, featuring 33 questions amongst undergraduate medical students to understand their knowledge, attitude, and preparedness towards the Covid-19 pandemic.

## Methodology

An anonymized eSurvey developed on an online cloud-based website (Survey Monkey®) covered different aspects pertaining to the Covid-19, including the pathobiology, clinical features, management, outcomes, impact on academics, and prevalent concerns regarding the same (supplementary table 1).

## Design of the questionnaire

Overall, the questionnaire featured 33 questions, most of which were multichoice. While three items were to identify respondent characteristics, 13 and 17 belonged to the factual and opinion set, respectively. The Likert scale was used to record responses in the opinion set.

The average time taken by each individual to complete the survey was eight minutes. The respondents could change the answers before submission, but not after it. All questions were mandatory. Internet Protocol address checks were done to avoid duplicated responses from a single respondent. Three professors and one undergraduate medical student reviewed the questions and confirmed them to be representative of the content validity of the survey. The survey was filled three times to check for errors in wording, grammar, or syntax. Correct responses were obtained from the Centre for Disease Control and World Health Organisation.^2,3^.

## Student selection

The questionnaire was served to undergraduate medical students in six medical colleges across the country. The survey was circulated over the email and WhatsApp® groups, primarily among 3rd MBBS part two students and interns. The eligible participants were given a week’s time to voluntarily complete the questionnaire from 26th March to 2nd April 2020. Informed consent was obtained at the beginning of the survey, and no incentives were offered for survey completion.

An exemption from review was obtained from the institute ethics committee of Sanjay Gandhi Postgraduate Institute of Medical Sciences, Lucknow, as per local guidelines.^4^ We adhered to the Checklist for Reporting Results of Internet E-surveys to report the data.^5^ Descriptive statistics were used, and figures downloaded from surveymonkey.com®.

## Results

Of 2500 invitees across six medical schools in India, 616 (24.6%) responded. Most undergraduates (age 21.5 years, 46.1% males) had recently completed (16.7%) or in the final year of their undergraduate training program (54.1%). Knowledge regarding the viral transmission 606 (97.7%), laboratory diagnosis 536 (87%), and drugs being tried out to treat Covid-19, 585 (95%), was adequate among most students (Supplementary Table 1). However, the understanding of the incubation period 123 (20%) and time to symptoms 30 (4.8%) was less than satisfactory.

Three-fourths (75%) were not aware of the treatment guidelines for Covid-19 and one-quarter (155, 25.1%) were unaware of the precautions needed while managing patients with the disease (Supplementary Table 2). Moreover, 179 (29.1%) were unaware that Covid-19 causes an asymptomatic or minor illness in most young individuals.

While the universities had advised most regarding hand hygiene, social distancing, symptom identification, and high-risk groups, fewer were advised to avoid staying back in hostels or drug prophylaxis (Table 1). More than half of the respondents (54%) either didn’t have (29%) access to N95 masks or have access occasionally (25%).

Nearly 20% were not sure if they had been in contact with or cared for someone with Covid-19 in the prior two weeks, and another 10% continued to attend clinical rotations, ignoring their symptoms suggestive of Covid-19.

Moreover, three-quarters (427, 69.3%) expressed reluctance to attend clinics from fear of getting infected or passing the infection on to others. Besides, 250 (40.6%) were not updated on Covid-19, and most (486, 78.9%) resorted to social media for information on Covid-19 (Figure 1).

**Figure 1.**
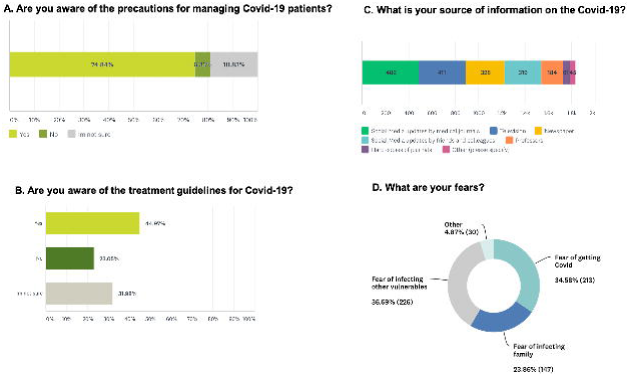
Knowledge and prevalent fears pertaining to the pandemic.

## Discussion

An increasing role of medical students in spearheading a voluntary task force while gaining skills and experience has been proposed in recent times.^6^ However, this survey identifies a knowledge deficit and insufficient awareness of the preventive and treatment strategies amid poor reading practices among students. An alarming 50% did not have access to masks, and one fifth were uncertain regarding contact with Covid-19 suspects. Few continued to attend clinical rotations despite having symptoms suggestive of active infection with the virus. This raises serious concerns regarding the knowledge base and flawed attitude of the medical students towards the ongoing pandemic.

Most suggestions to recruit students into the healthcare workforce stem primarily from the relatively benign course of Covid-19 in those aged below 40 years. ^7^ While students were adept with the clinical features and diagnosis, the knowledge base was inadequate regarding the incubation and time to symptoms, which is vital to advise precautions, especially the quarantine duration. Additionally, ignorance and lack of concern about the pandemic’s seriousness were reflected in their attitude, as they continued to attend clinical rotations despite having symptoms suggestive of Covid-19. Moreover, the inability to identify if they have had exposure to Covid-19 patients raises a grave issue of knowledge deficit and poor attitude. The Medical Council of India and various medical colleges need to make serious efforts to educate undergraduate students on an urgent basis through online lectures and training modules. The government agencies need to factor in that the undergraduate medical students are either at similar risk or more to get infected as the other HCWs if they are made to serve as front-line team against Covid-19 in India. Additionally, they are not licensed to practice before the mandatory internship rotations hence lacking in the requisite skillset for serving the. Covid-19 patients during the pandemic Any undue haste on the part of government officials may thus lead to disastrous consequences. Although the unusual circumstances have forced government agencies to expedite registration and practice in certain countries such as the United Kingdom, the outcomes of such actions require close monitoring. ^8^

In the worst-case scenario where there is a severe shortage of HCWs and government is forced to utilize the services of undergraduate medical students, the following could be the best way of their utilization; care of non-Covid patients under supervision, assistance to senior doctors, part of screening services, coordination of interdepartmental efforts, and public outreach and community education programs. The advantages of this would be manifold, from maintaining the continuity of medical training, helping students develop a competency-based skill base, and, most importantly, gather the unique experience of serving the community in a pandemic, that would last them a lifetime. However, the identified deficits call for significant education and training to improve upon their existing knowledge bank before they can embark on such endeavors. It is essential to be mindful of the potential harms of misinformation, especially while dealing with the masses through public outreach programs. It is also essential to understand that any participation in medical undergraduates should only be voluntary.

The prolong lockdown in various countries has led to a visible shift amongst doctors to social media platforms for updates on scientific literature^9^, and medical students are no exception. Although the identified deficits in the knowledge base can be honed by online training, the ensuing infodemic has made it challenging to locate credible information in the haystack.^10^ Resorting to sources (local guidelines issued by scientific agencies) that provide verified information assumes even greater importance at such times, especially for care providers. Indian council of medical research, a government agency, is regularly publishing updated guidelines on Covid-19, and medical universities should circulate among students.^11^

The limitations of our survey are less than expected (25%) response rate (ideal response rate 60% or more) and the inherent biases of self-reported questionnaires. This could be due to the short (week-long) duration of the survey and request over one social media platform. Moreover, time is of the essence in understanding the attitude of students towards the pandemic. Since this is a policy issue that deserves quick and urgent attention due to the unusual situation, apriori, a cut-off of 500 responses were deemed appropriate. This survey is the first study evaluating the knowledge and attitude among undergraduate students, providing a deeper understanding of their preparedness against the ongoing pandemic.

Thus, we conclude that inadequate understanding of Covid-19 and prevalent fears amongst young medical undergraduates found them under prepared for Covid-19 care. If their services are to be utilized during the pandemic, focused education should first be imparted. This survey calls for serious introspection by health authorities before contemplating such action.

## Data Availability

All data collected through surveymonkey

---

Supplementary Table 1. Knowledge base in relation to Covid-19

Supplementary Table 2. Attitude of medical undergraduates regarding Covid-19

